# Development and exploratory analysis of a multi-dimensional metric of adherence for digital health interventions

**DOI:** 10.1101/2024.02.23.24303246

**Authors:** Harry T. Mason, Siobhán O’Connor, David Wong, Emma Stanmore

## Abstract

**Introduction:** Adherence is often cited as an important metric to demonstrate sustained engagement of an individual or population with a health technology, but its definition is often ill-defined. Any adherence definition made for digital health interventions must be clearly defined to ensure a consistent approach to measuring sustained use as an indicator of impact.

**Methods:** This study followed mathematically-defined definitions of distinct aspects of adherence: initial adoption, consistency, duration, and dropout. These were then applied to a digital physiotherapy dataset of older adults (N=56). Participants were assigned 3 sessions a week of exergames (exercise-based videogames) for 12 weeks using MIRA rehab software platform.

**Results:** The following adherence characteristics emerged: an initial dropout of 3% (completed ≤3 sessions), 20% of participants achieving the desired consistency (≥3 sessions a week for 12 weeks), 39% of participants passing a duration threshold (completing ≥20 minutes a week for 12 weeks), and an average dropout at 72.3% (when judged by percentage of sessions completed at dropout).

**Conclusion:** The approach used for measuring and reporting adherence metrics allows readers to draw clear conclusions about the different aspects of engagement that users displayed with the digital health programme. This type of reporting is recommended for all future digital health studies reporting adherence measures to ensure a consistent approach to reporting and comparing digital health interventions and their impact.

## 1. Introduction/Background

The use of technology in healthcare (known as digital health) has proliferated over recent years due to their potential in enhancing efficiency, responsiveness, personalisation, and ability to embed behaviour change techniques to improve outcomes. Digital health encapsulates the intersection between healthcare and emerging or established technologies^1^ and include mobile health, wearable devices and telehealth. A prominent example of an emerging technology is gamified telerehabilitation, also known as exergames^2^. Exergames are active video games that use body movements in physical exercise as virtual controls within the gameplay. Exergames provide opportunities for home-based rehabilitation training and longitudinal participant monitoring^3,4^ that would be costly to achieve with traditional physiotherapy.

One common concept used to define the success or failure of digital health approaches is “adherence”. There are multiple ways in which adherence can be defined, all of which encapsulate one of more aspects of engagement a user has with this new approach^5^. Demonstration of adherence is a necessary causal precursor to any clinical outcomes^6^. In the context of exergames, adherence is of particular interest as a measure of increased or decreased use through the instantaneous feedback that exergames are expected to provide. In addition, the gamification elements of exergames (such as points and level ups) may provide additional incentives^7^ to complete a prescribed programme (to ensure sufficient dose, challenge and progression) that would not be present in traditional physiotherapy.

Despite the promising performance of exergames in different participant groups (e.g., older adults^8,9^, and children and adolescents^10,11^), definitions of adherence vary, and relatively few studies compare adherence in exergames to other forms ofexercise^2^. Exergames fall at the intersection of healthcare, exercise, and technology, all of which typically have different motives when defining adherence. The World Health Organisation (WHO) definition of adherence focuses on provider-defined change (“the extent to which a person’s behaviour – taking medication, following a diet, and/or executing lifestyle changes, corresponds with agreed recommendations from a health care provider”)^12^. Technological approaches may focus instead on usage, and the distinction between usage and other aspects of engagement in digital Health can be inconsistent^13^. Different terms for adherence in separate fields (such as concordance or compliance) only serves to further obfuscate matters.

Skjæret et al.’s systematic general review of exergames^2^ found that, of the studies they investigated (n=54), over 10% did not report the number of participants that completed the study. Almost half did not report on the completed sessions during the intervention, and for the rest the adherence methods varied between reporting raw number of sessions completed, the percentage of sessions completed, and the degree of accuracy with the prescribed intervention. Many of these descriptions of adherence may prove useful, but the inconsistency in their meaning and measurement makes it hard to draw any systematic conclusions about how to define and assess this important concept.

From a user perspective, monitoring adherence is also important as it can enable a digital tool to provide useful feedback in terms of how often and how well someone follows an exercise regime, or not, and give tailored advice that could improve health outcomes long-term. From a service perspective, adherence data is also valuable to help plan and manage the demand for digital health services locally and nationally and how these can be adequately resourced and delivered. From a research perspective, assessing adherence can be beneficial and viewed at either a personal level or a population level. The personal level (e.g., comparing completed vs. assigned sessions of exercise at any given time) may have practical purposes of real time adjustment of workload or predicting dropout. At the macro-level, analysing adherence data could be useful for improving system/technology design and drawing conclusions about the overall effectiveness of research studies on telerehabilitation.

Regardless of the individual or organisation monitoring adherence, the term and its measures should be applied consistently. Useful dimensions of adherence relevant to digital health should be separated from dimensions of adherence related to other types of interventions which are more universally calculable. Furthermore, conclusions in one study drawn based on adherence can only be compared with another study if the same core underlying definition of adherence is clearly defined and agreed upon.

Here, existing scientific literature is used to define a set of individual and common dimensions of adherence relevant to digital physiotherapy interventions (telerehabilitation). Then a set of metrics to quantify each dimension is proposed to provide a holistic set of metrics for adherence. This set of metrics is applied on a digital physiotherapy dataset and exemplar conclusions are drawn about what these different adherence metrics tell us about this dataset, and to provide a basis for future adherence reporting.

## 2. Methodology

### 2.1 Study design

A secondary analysis of a dataset from a clinical trial of a digital physiotherapy intervention was undertaken.

### 2.2 Ethics

This research complies with the Declaration of Helsinki. Ethics approvals for this study were obtained from London - Camden & Kings Cross Research Ethics Committee, UK, reference number 16/LO/0200. All participants provided written informed consent prior to entering the study. The trial was registered at ClinicalTrials.gov on 18 Dec 2015 with reference number NCT02634736.

### 2.3 Dataset

An anonymised dataset of adult participants (n=56), age 58-96 (mean age =78, 45 female), were acquired from two clinical trial field sites in Manchester, United Kingdom (UK) (n=41) and Glasgow, UK (n=15) in 2016. The digital dataset^14^ was gathered in collaboration with MIRA (Medical Interactive Rehabilitation Assistant), a computer software package designed by MIRA Rehab Limited^15^ that gamifies physiotherapy exercises using motion tracking and active video-games. Each participant was assigned several “games“, to make up a programme for each exercise “session“. This may be considered an exergame equivalent to an in-person physiotherapy session made up of different exercises. Exercise sessions in this dataset were deliberately designed to be short to match the pre-frail to frail level of ability for older adults with multiple health conditions recruited from Assisted Living Facilities. Each participant was then asked to complete a minimum of three exercise sessions a week, for a series of 12 weeks. Duration of exercise and date of session completion were automatically recorded. The dataset included the type of digital games that were completed, but not the initial list of games that were assigned to each participant.

### 2.4 Measuring Adherence

Five dimensions of adherence – initial adoption, consistency, duration, dropout, and intensity - are defined here in the context of this type of study. They are well-defined in contexts for which repeated interventions are required, for instance, in randomised controlled trials of pharmaceutical interventions^16,17^. These separate dimensions also capture many of the different ways in which adherence is described in the exergame literature^2,4^. Metrics were developed based on these five dimensions. *a_subcategory_* refers to an individual’s normalised adherence score for that dimension subcategory. The population-level analysis can then be derived from the trends observed in individuals.

#### 2.4.1. Adherence as initial adoption

The first adherence metric is designed to separate users into two groups depending on whether an individual passed an initial engagement threshold for the new digital physiotherapy interventions. This metric could also be considered to measure the amount of people that pass the “curiosity plateau”, that is, how many people keep using the digital health programme once the initial novelty of using it has worn off. This adherence metric (a_adoption_) is defined for a given program by how many sessions a participant completes in the recommended timeframe (s), and whether that surpasses a defined threshold (ɵ_s_). Due to the study ascribing three sessions a week, ɵ_s_=4 was set to represent a participant doing at least a week’s equivalence in sessions (Equation 1). Additionally, a “session days” metric was included - the days in which at least one session of digitally-supported exercise occurred. This metric will help distinguish between those who exercised three times a week across the week, compared to those who completed three sessions in a single day (with s referring to “session days” instead of “sessions” in Equation 1). Duration and amount of exercise are not captured in this metric.

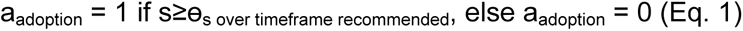

#### 2.4.2. Adherence as consistency

Another useful dimension of adherence is the consistency with which a participant undertakes at least a certain amount of exercise with respect to given timeframe. Here, the number of sessions completed per week (w_s_) must at least equal a threshold of three sessions per week (ɵ_w_) across the entire 12-week period (N_w_). Where a participant drops out of the digital physiotherapy intervention before the end of their recommended plan, only consistency during pre-dropout weeks are considered. This metric is considered useful to evaluate whether habit-forming behaviour has occurred^18,19^.

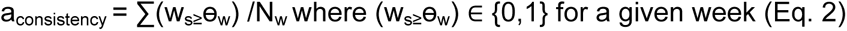

#### 2.4.3 Adherence as duration of exercise

Adherence as duration is defined by whether a participant undertakes at least a certain duration of exercise with respect to given timeframe. Here, the duration of exercise completed per week (d_s_) must surpass either 20 or 30 minutes per week (ɵ_d_) across the entire 12-week period (N_w_). The 20- and 30-minute thresholds were both considered separately, selected as representative amounts of exercise within the context of the MIRA exergame interface. Like the consistency measure, this establishes whether an individual was assigned a plan appropriate to their personal levels of ability to complete it. Where a participant drops out of the digital physiotherapy intervention before the end of their recommended plan, only duration during pre-dropout weeks are considered.

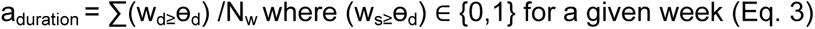

#### 2.4.4 Adherence as dropout

Another useful metric to consider is the rate of dropout over the course of a prescribed digital exercise plan. The Kaplan-Meier curve (and the area under the curve) measures the proportion of a plan completed by an individual, and serves as a well-established metric for this calculation^20,21^. Kaplan-Meier provides a good indication of the dropout rates over a given time-period when considered as a full population, although it does require a full plan to be pre-defined (i.e., without the flexibility for additional exercises at the end of an initial batch of sessions). The traditional Kaplan-Meier graph measures the time engaged in a plan (t_completed_, the time between the first and last recorded session) compared to the total length of the plan (t_recommended_, i.e. 12 weeks), which does not provide any information about how consistent/active a participant was in a plan before they dropped out (e.g., a participant doing one exercise a week for five weeks looks worse than one doing the full plan for four weeks). As such, a time-independent version of Kaplan-Meier is also included using number of sessions completed (s_completed_) compared to the total recommended (s_recommended_, e.g. 36 sessions), which measures percentage the completion of the digital exercise plan achieved when the participant drops out. a_dropout_ is not allowed to exceed 1.

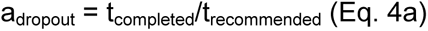

or

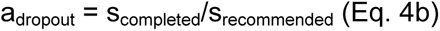

#### 2.4.5. Adherence as intensity

Average exercise intensity is an additional measure sometimes considered in the digital health literature^21,22^. While the “scores” for each were accessible exergame (indicating an objective measure of physical achievement), this metric was not considered to have a natural equivalent to non-exergame programmes. As such, this metric will not be considered in this study. This is the most subjective of the adherence measures, and so care must be taken when designing a metric for this purpose in the future. Using the first four adherence metrics (i.e., initial adoption, consistency, duration of exercise, and dropout), the clinical trial data were analysed to produce a set of adherence metrics for the digital physiotherapy intervention that was provided to participants.

## 3. Results

Figure 1 shows how adherence as initial adoption varies depending on the base threshold used (ɵ_s_). The figure shows that one participant only completed 3 sessions, and a second participant completed only 4 sessions instead of the full 36 sessions. All other participants completed at least 8 sessions. However, the lower graph shows that a third person only completed at least 3 “session days” – meaning they were highly active during the days they did try the system but did not stick with the programme beyond those days. Different digital physiotherapy interventions may find differing thresholds and measures to be useful here.

**Figure 1:**
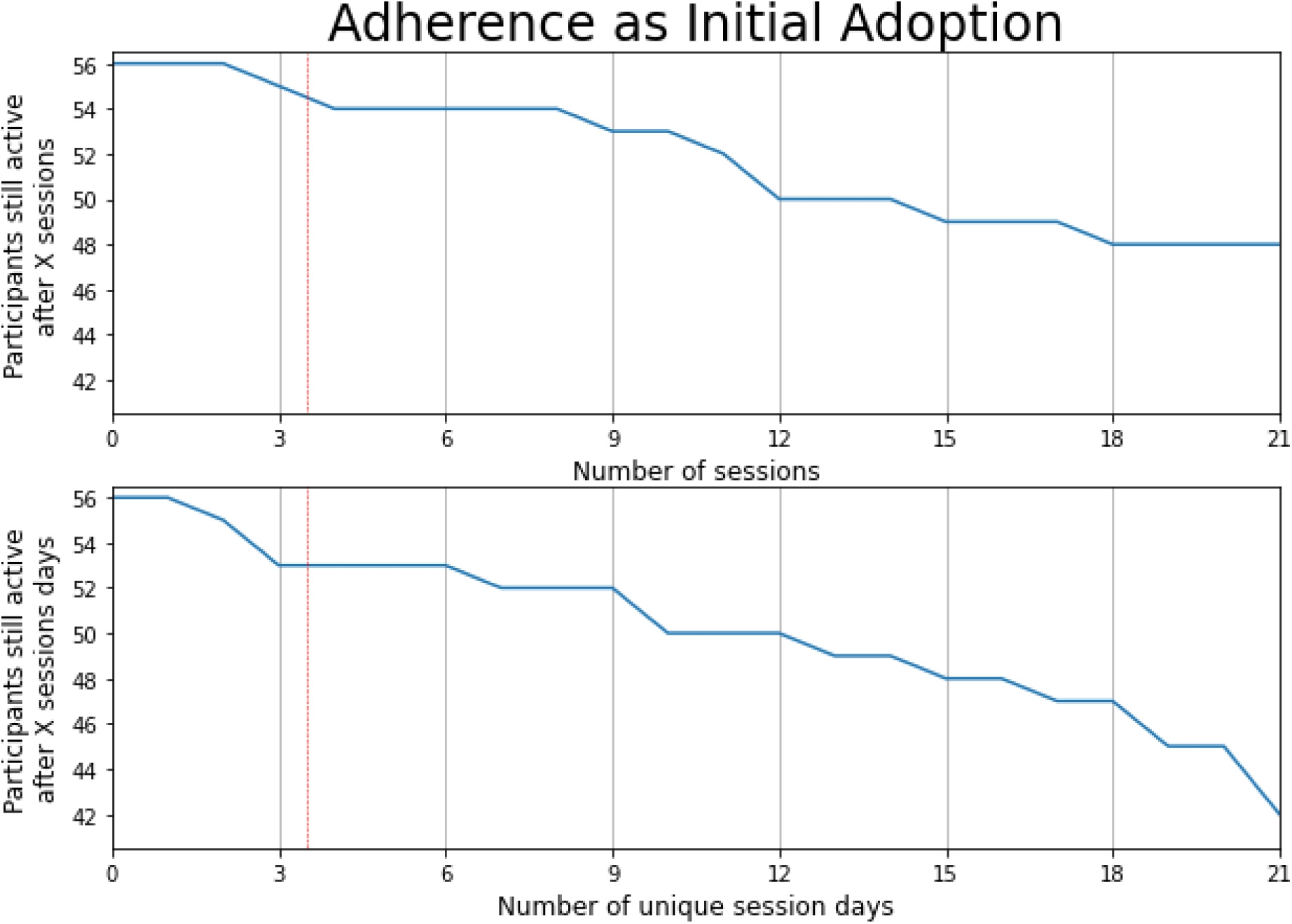
A figure showing the number of participants who are still active in the clinical trial, given a certain number of (top): sessions or (bottom) unique sessions days. The study started with 56 participants. Participants were asked to complete 3 sessions a week. The dotted line represents ɵ_s,_ in equation 1 the threshold used to define initial adoption vs initial non-adoption. As this metric is focussed on initial adoption, only the effect of the threshold ≤21 days is shown here.

Figure 2 shows the consistency measure applied to the trial dataset to gauge this dimension of adherence (Eq. 2). Here, only a full week of completed sessions (recommended as ≥3 sessions for that week) is considered in the analysis. The number of ≥3 session weeks is then tallied for each participant. This graph is used to define how many participants consistently adhered to their recommended plan. All participants beyond the red line completed all 36 sessions at the recommended level of exercise. 45/56 participants (80%) failed to complete ≥3 sessions a week for 12 weeks. Some participants made additional recordings on the digital physiotherapy intervention once their initial 12 weeks were completed. The top subfigure is useful for identifying common stopping points within the programme (e.g., a considerable number stopped after achieving only five full weeks, just short of halfway goal of the programme). The bottom subfigure is useful for identifying the number of participants that achieved a certain consistency metric (e.g. 8 participants failed to complete ≥3 sessions a week for at least 2 weeks).

**Figure 2:**
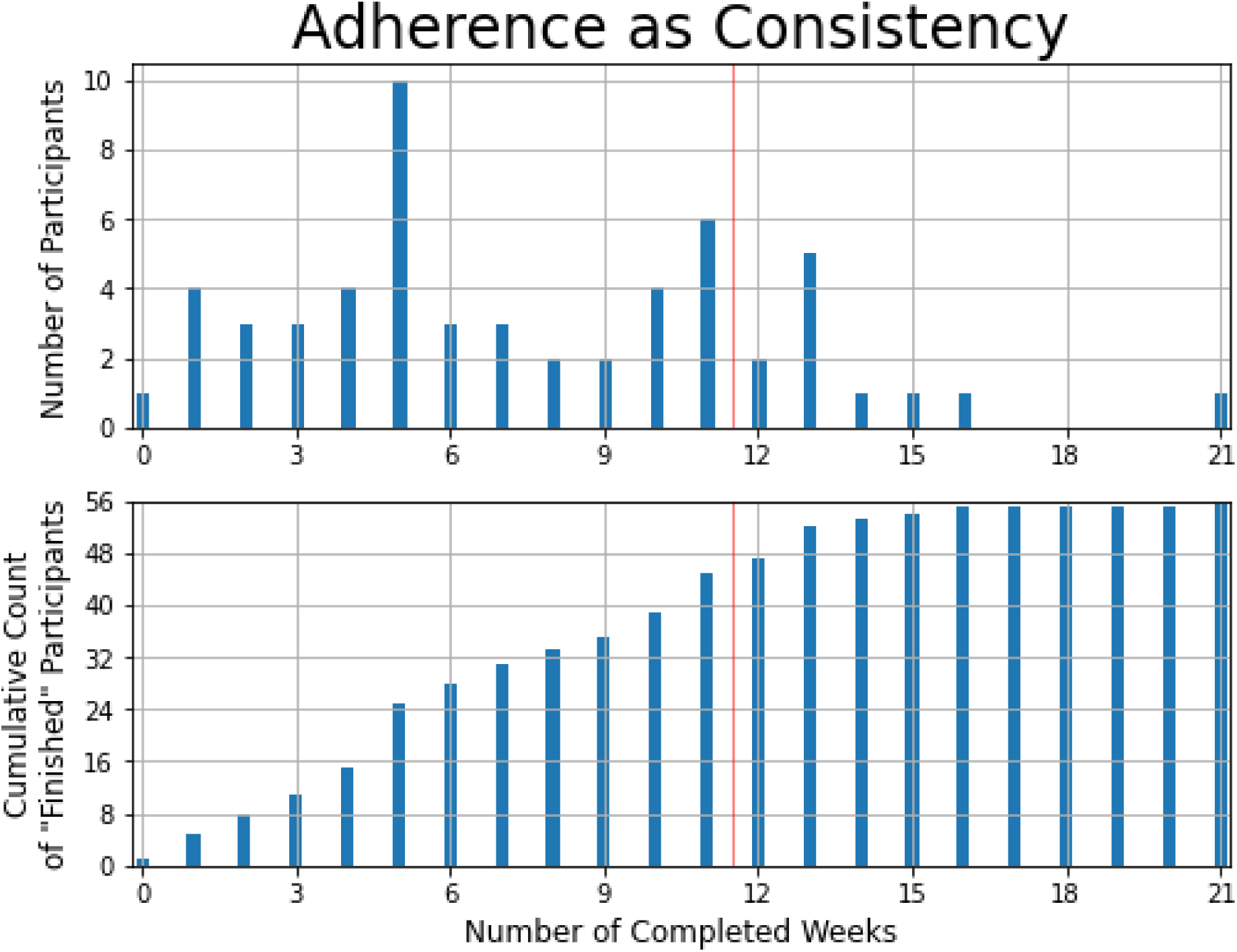
The number of participants that achieved ≥ 3 sessions a week for N weeks, as defined by Eq. 2. The thin red vertical line at 11.5 weeks indicates that all the participants beyond that line completed the recommended exercise plan. Some participant data is recorded beyond the end of the study. The top graph shows the distribution of total weeks completed; the bottom graph indicates the cumulative number of participants who failed to achieve a consistency threshold for specific number of weeks.

Figure 3 demonstrates how duration of exercise can be used as an adherence metric (Eq. 3). Similar to the consistency measure, an activity threshold is used to define an appropriate level of engagement over a timeframe. Here, only weeks in which a participant achieved either 20 minutes per week or 30 minutes per week (for the left and right subplots respectively) are considered, as this was the recommended dose of the digital physiotherapy intervention. 34/56 participants (61%) failed to complete at least 20 minutes per week for 12 weeks, as opposed to 43/56 (77%) of participants who failed to complete 30 minutes per week over the same period. The figure can be used to help determine how representative these results are (e.g. 3 participants reached the threshold for 11/12 weeks for both thresholds but would not be represented in a final summative report).

**Figure 3:**
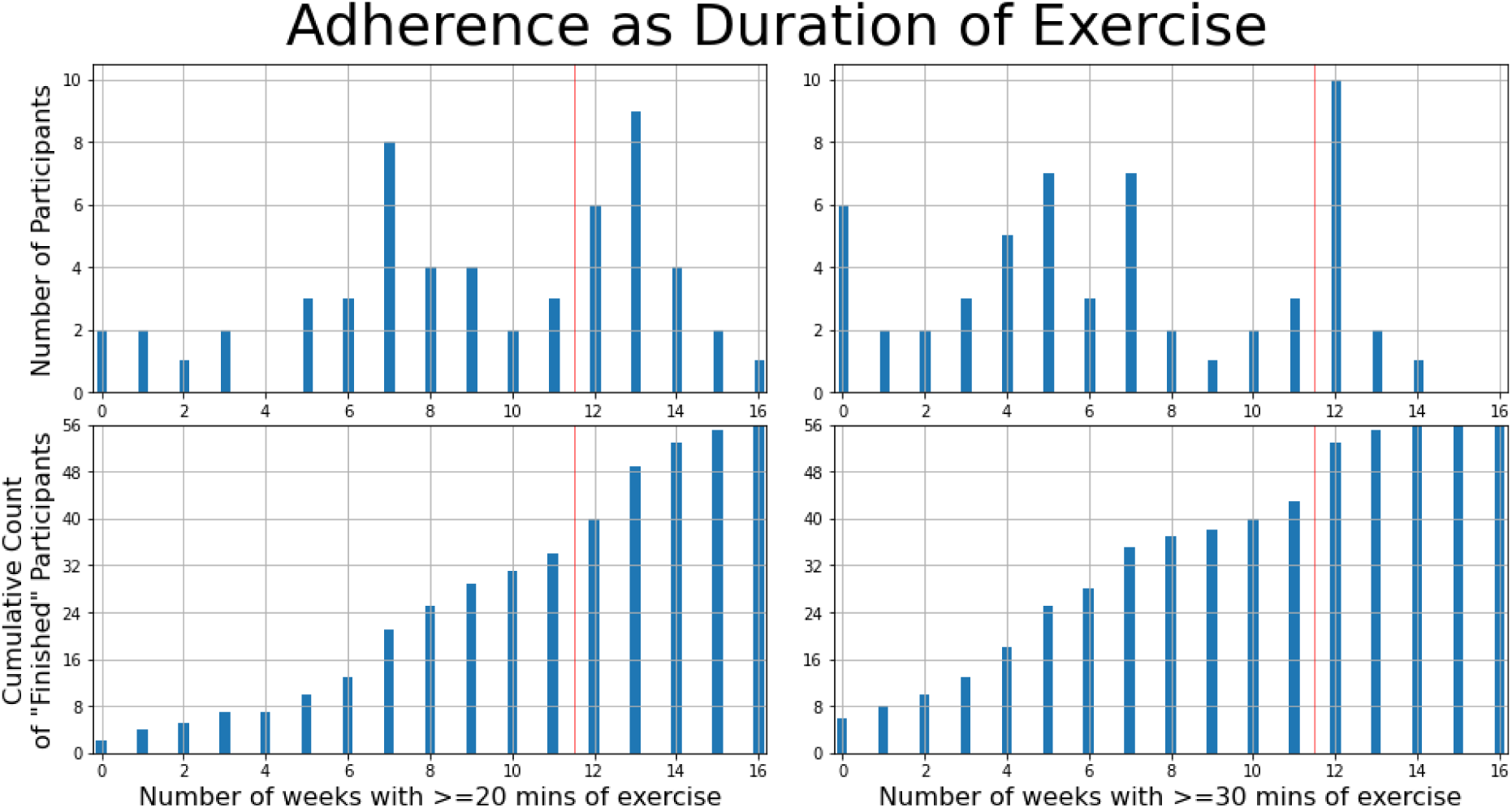
The number of participants that achieved ≥20 (left graphs) or ≥30 (right graphs) minutes a week for N weeks (Eq. 3). The thin red vertical line at 11.5 weeks indicates the recommended study length. Some participant data is recorded beyond the end of the recommended exercise plan. The top graphs show the distribution of total weeks completed; the bottom graphs indicate the cumulative number of participants who did not meet the duration threshold for a given number of weeks.

The adherence as dropout metric is displayed in figure 4. The top graph mimics the Kaplan-Meier graph used in clinical trials, displaying the use of the telerehabilitation system by the length of time that any participant was still involved in the study. It does not indicate the consistency or level of exercise during the programme. It serves as a more naturally interpretable visualisation of dropout, with a high initial compliance followed by a steeper decline after 75 days. It is worth noting that 84 days would have been the natural stopping point for any individual who completed the full 12 weeks, although the nature of this dataset means that records of many individuals who completed further sessions were available for analysis. Although 84 days was the initial cut-off, it is still worth noting that 80% of individuals made it past the 75-day mark. The bottom graph of figure 4 measures the percentage of sessions completed, out of a theoretical maximum of 36. Any sessions completed beyond the initial 36 are ignored. The bottom graph more clearly indicates the progress through a programme for each participant but does not encode any time information. The AUC of the bottom graph is 0.723, where an AUC of 1 would represent zero dropouts throughout the duration of the study.

**Figure 4:**
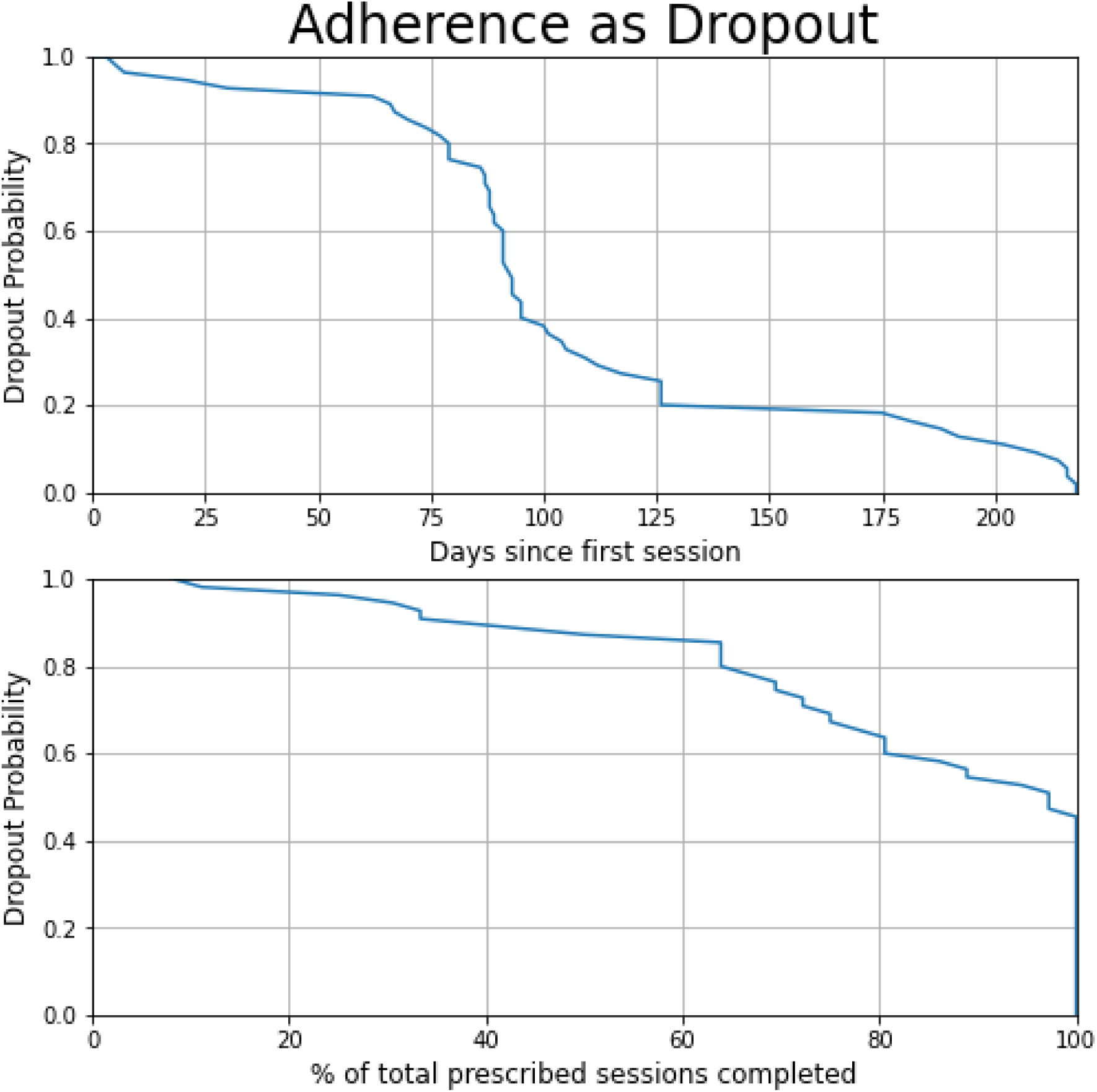
Adapted Kaplan-Meier plots, showing the rate of dropout (called “survival” in traditional Kaplan-Meier) by participants across the recommended digital exercise programme (Eq. 4). Top: the number of days between the first and the last session; Bottom: The % of sessions completed out of the recommended 36.

Figure 5 shows the relationship between the separate dimensions of adherence. Initial adoption serves as the simplest metric, with the two exclusions also having low consistency and duration of exercise. The 30-minute duration threshold shows lower visible correlation with the consistency and dropout metrics than the 20-minute duration threshold. While correlation for the consistency, 20-minute duration, and dropout metrics is high it is not perfect, indicating that these dimensions are capturing the same underlying data, but capturing slightly different perspectives of when a given participant may stop adhering to the study protocol.

**Figure 5:**
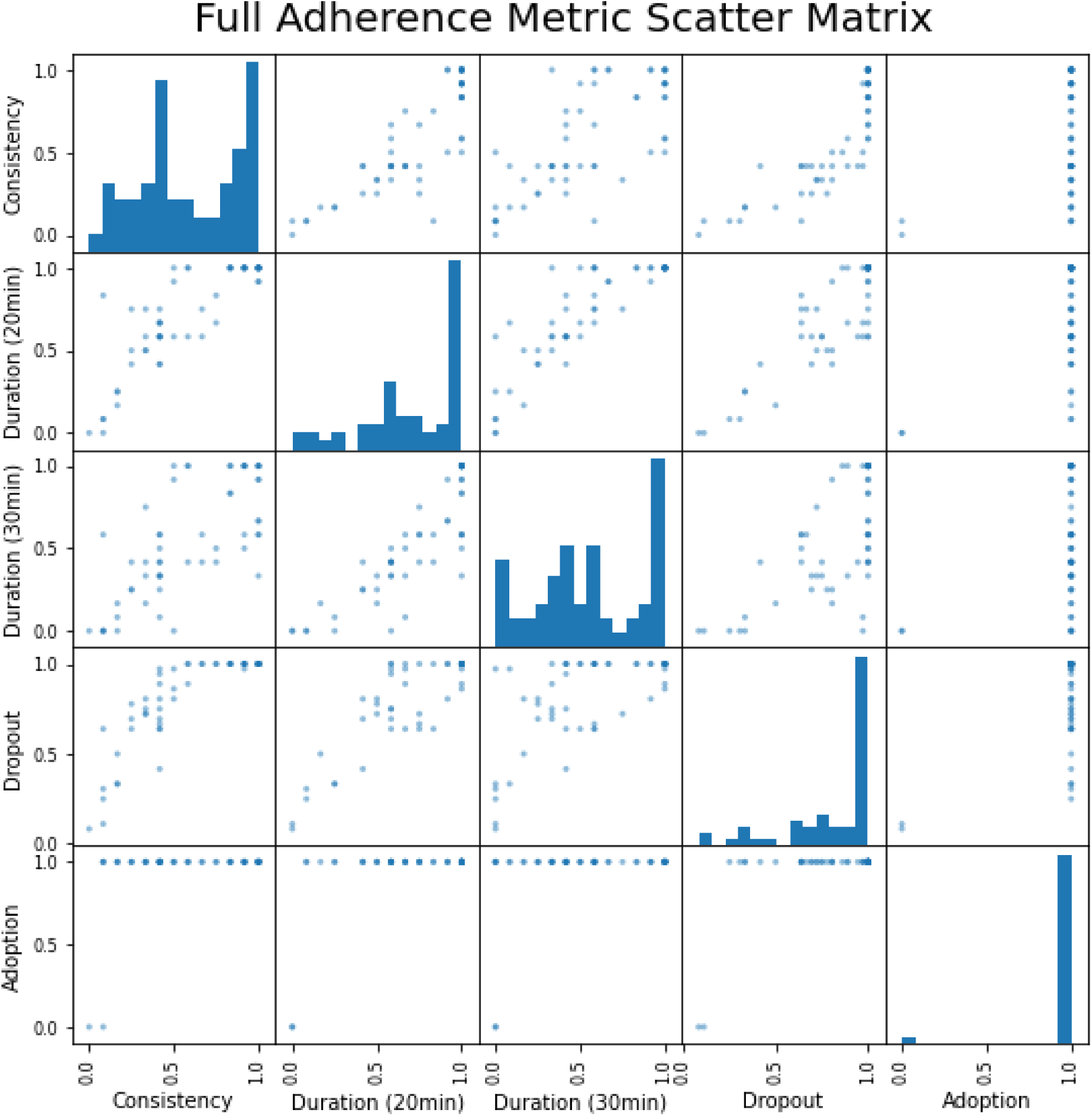
The relationship between adherence metrics. Each participant is assigned a normalised score for each metric. The diagonals show the score distribution for each metric, and the off-diagonals show the pairwise relationship.

While group population metrics are vital to understanding the overall adherence to a digital physiotherapy intervention, it is also important to visualise exercise curves for each participant so individual level adherence can be gauged. Figure 6 shows four participants who all did less than 36 sessions, but who all had quite different progression curves. Participant 1 completed 24 sessions in 9 weeks (2.7 sessions/week average), with a clear gap around week 7. Participant 2 only missed one session in week 9, and otherwise completed 35 sessions in 12 weeks. Additionally, Participant 2 completed at least 35 minutes of exercise each week while also increasing their total weekly duration throughout the study. Participant 3 had a long gap around weeks 9 and 10, but otherwise had a similar average completion to Participant 2. Participant 4 had a long duration of exercise, with two separate one-week gaps early in the study. These data could be provided to each participant so they understand their daily and weekly exercise progress and personalised recommendations based on these individual data could be developed and provided via electronic notifications so participants can improve their fitness regime.

**Figure 6:**
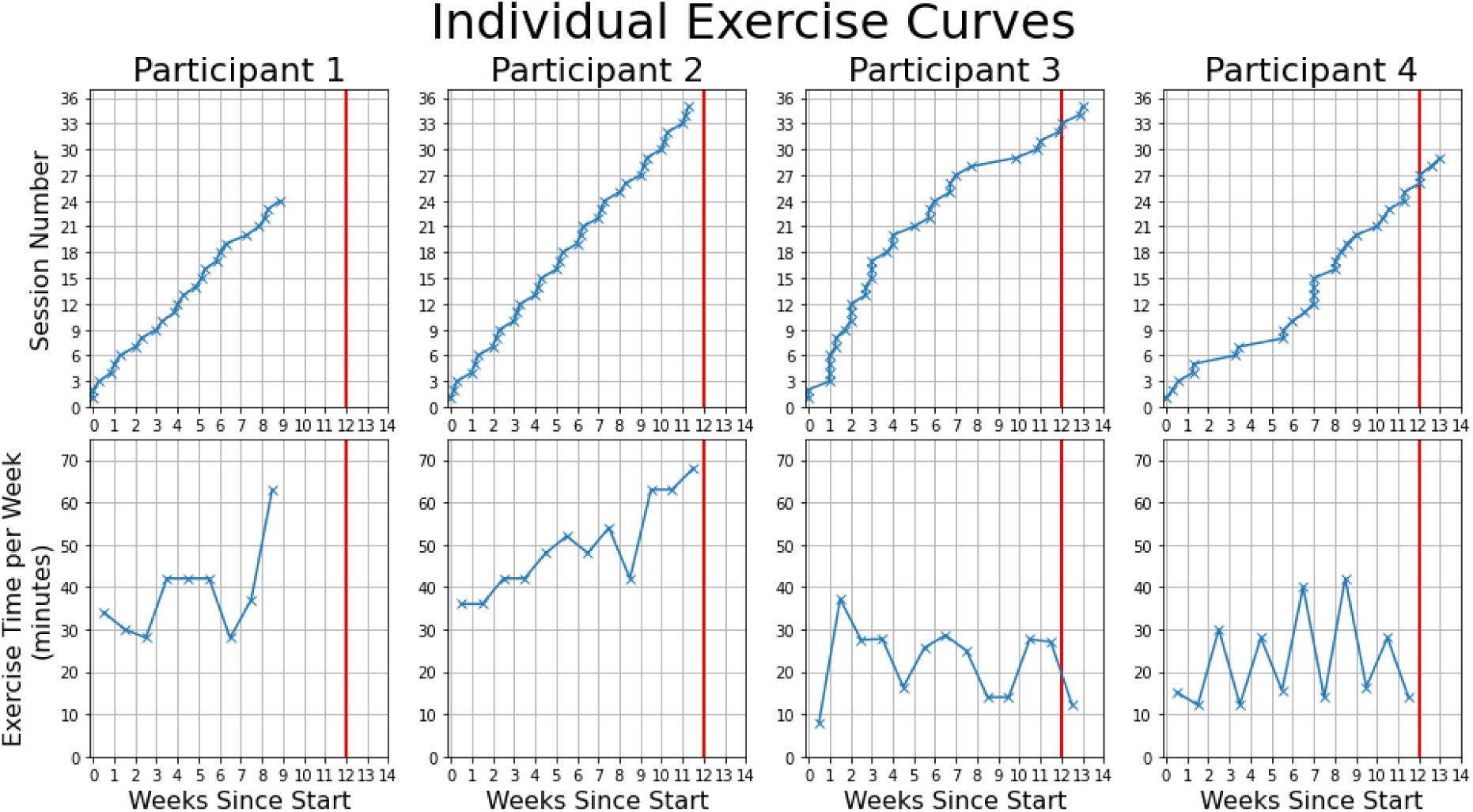
The consistency and exercise duration metrics of a subset of participants in the study. The red line indicates the 12-week line, by which an “ideal” participant would have completed 36 sessions.

## 4. Discussion

The secondary analysis of the trial dataset has enabled a more in-depth exploration and development of key adherence metrics for a digital physiotherapy intervention.

The choice of ɵ_s_ = 4 sessions per week serves as a useful cut-off point for initial adoption (i.e., accepting participants who completed over an equivalent week of recommended sessions) in relation to this intervention, but other baseline usage and timeframes may be appropriate for other types of digital health interventions. Two participants were dissuaded from continuing from this point onwards, yielding a 3% rate of initial dropout. All other participants continued until nearly the 3-week mark or further. These participants have not been excluded from any subsequent measures to allow for cross-dimensional analysis.

Some reasons for dropout are listed for this dataset by Stanmore et al.^14^, with some motivations being potentially linked to the intervention (e.g. disinterest with technology), while some are separate (e.g. becoming medically unfit or family issues).

The consistency and duration of exercise are certainly linked (Fig. 5), although they are still worth considering separately. The habit-forming nature of regular exercise is a vital indicator of long-term success, especially in the case of physiotherapy^23,24^. Adherence as duration is a marker of the quantity of exercise and might be much better to pair with any physical outcomes (e.g., increased range of movement). It is worth noting that this study did not assign a fixed duration of exercise to the participants, which is likely why the ≥30-minute threshold of Fig.5 produced such spread results when compared with consistency. When considering the full plan only 20% of participants successfully completed ≥3 sessions for 12 weeks (median: 5.5 weeks), only 23% completed ≥30 minutes of exercise a week (median: 5.5 weeks), and 39% completed ≥20 minutes of exercise a week (median: 9 weeks).

“Adherence as duration” is particularly critical if analysing outcomes such as falls for preventative programmes^4^, which suggest 180 minutes/week of progressive exercise for the greatest effects^25^. Conversely, an older adult is advised to complete 150 minutes/week of exercise according to physical activity guidelines^26^, and so it should be noted whether an exercise plan is given above and beyond this baseline amount. “Adherence as consistency” can be useful in considering target implementation programmes, tailoring a programme to best fit a participant’s personal routines.

The dropout metric is a much more useful gauge of a whole study, and it would be interesting to observe in future studies how the curves change for different digital health programmes. There were two separate plots used for dropout for this study (Fig. 4). In many cases, the sessions will be of a non-fixed duration, and so the time-based measure of dropout will have to be used. Where the digital exercise programme is more structured, it is likely that the percentage-completion metric will prove more useful to researchers. The time-based dropout measure showed that over 80% of participants had at least 5 days between their first and last recorded session. The AUC of the percentage-completion graph was 0.723, which could be interpreted as 72.3% of the assigned sessions were completed (discounting additional sessions completed beyond the 36). It is also interesting to note that, despite those who completed the 36 sessions all having an a_dropout_ score of 1, there were a range of consistency and duration values amongst that subgroup (Fig. 5).

The four dimensions – initial adoption, consistency, duration, and dropout - all provide distinct information on the adherence to a digital health intervention, while also identifying specific areas where improvements may be required to match the recommendations of the programme. A low initial adoption/usage rate could indicate an overly complicated software interface or hardware component, poor Internet access, and/or low levels of digital literacy, among other issues that prevent people using digital health tools^27^. A low consistency could be improved with a reminder system. A low duration of exercise might be addressed with reducing the intensity. If the dropout is extremely high at a given point, it might be worth observing whether the digital exercise programme introduces a difficult exercise at that point.

While it is possible to reduce each of these adherence dimensions to a single summative statement, it is important to note the nuance lost in doing so. Figure 6 shows four participants that each failed to complete the full 36 sessions. Participant 2 missed a single session, and to all intents would likely have very similar outcomes to a participant who completed the full programme. Participant 3 also essentially completed all sessions, but did so over a slightly longer time period, which may be a confounding factor for some programmes depending on the degree of time extension. Both participants completed a much greater percentage of exercise than Participants 1 and 4, and so it may be useful to consider adherence as a more continuous metric that would encapsulate these variations.

Evaluating the efficacy of the trial by comparing the level of improvement between those who completed the assigned set of exercise and those who did not was considered beyond the scope of this paper. It is noted that the duration and consistency metrics could prove an interesting variable to evaluate such outcomes, and that such an analysis would remove the issue of reducing adherence to a single metric. The use of population adherence metrics and individual adherence metrics provide a chance to evaluate overall study effectiveness, as well as identifying individual behaviours within a study. Additionally, examining how these metrics vary across demographics (e.g., age, gender, ethnicity, sexuality, level of education) could provide more precise information about characteristic adherence in separate populations. Finally, a metric designed to capture the “intensity” metric of adherence could also prove useful in efficacy calculations, provided it follows the same general principals established here (e.g., be able to be applied fairly between digital health and non-digital health datasets).

Here, separate adherence metrics have been developed to provide distinct insights into how participants use a given digital health intervention. While “sessions” were distinctly defined here via recommended guidelines, they could also be more broadly defined as “periods of continuous exercise” or “days containing exercise” for other interventions. It is important to note that there are areas where it is harder to reliably obtain information in non-virtual settings. Follow-up questionnaires are often used to gauge levels of exercise, which are more subject to drop out bias and require additional involvement from participants when compared to the automatic data collection of a digital tool. Hence, duration of exercise will also be far less precise in such circumstances.

### 4.1 Limitations

Global measures of adherence to digital health interventions were developed in this paper using metrics designed to be as broad as possible while still answering specific questions that allow digital health studies to have a non-digital equivalent. This paper applies four adherence metrics to one specific dataset, which will not capture the full nuances for all types of digital health interventions and will require further testing for consistency and test-to-test validation of the metrics. For example, a younger cohort would be assigned a greater duration of exercise per week, and the duration of exercise would have to be shifted accordingly. Additionally, numbers gathered automatically by a digital system are more accurate than the self-reported numbers of a non-digital equivalent. However, this an important starting point towards more informed analysis of the adherence to such interventions. While reported metrics will by their nature be tailored to the intervention in question, it is vital that all such tailoring is clearly stated to allow for accurate cross-study comparison.

In this paper, and in other studies, there will always be limitations enforced through the breadth of data captured. This can either affect the metrics of adherence directly, or alternatively the factors which affect adherence without being a direct metric. This paper is not designed to be an exhaustive list of all metrics that can be measured (e.g. a mobile health dataset can easily capture many more aspects of adherence than a standalone device^5^). Additionally, variable analysis can further correlate which factors are influencing distinct aspects of adherence (e.g. in this dataset, knowing which programmes were assigned to different individuals could provide feedback to engaging or non-engaging exergames).

Additionally, adherence is not the only useful metric that has been subject to unclear reporting. Integration of the adherence metrics with the TIDieR guidelines^6^ (specifically steps 8, 11, and 12) could help formalize the process and increase the quality of trial performance in future. Other related metrics (e.g. measuring participants who withdraw before a trial start date) could also prove useful to break down and integrate in future investigations.

## 5. Conclusion

In this paper, four complementary measures of adherence, initial adoption, consistency, duration of exercise, and dropout, to digital health interventions were developed and applied to an exergames dataset from a clinical trial. This dataset had low initial dropout (3%), but few participants completing the full course of the study (20% completed 3 sessions per week for 12 weeks, 39% completed 20 minutes per week for 12 weeks). Only 72.3% of the recommended exercise sessions were completed. Each of these metrics provides distinct information regarding the overall adherence to a digital physiotherapy intervention. The specific types of adherences evaluated are clearly explained and reported, allowing for a clear understanding of the fidelity of different aspects of the programme. The clear reporting of these metrics could prove useful in improving the relevant technology platform and user experience moving forwards.

## Data Availability

Data are available from the University of Manchester Institutional Data Access (contact via corresponding author Dr Emma Stanmore) for researchers who meet the criteria for access to confidential data.

